# Comparison of Macular Thickness Measurements Using Swept-Source and Spectral-Domain Optical Coherence Tomography in Healthy and Diabetic Subjects

**DOI:** 10.1101/2020.11.22.20236307

**Authors:** Kun Xiong, Wei Wang, Xia Gong, Wangting Li, Yuting Li, Jie Meng, Langhua Wang, Xiaoling Liang, Yizhi Liu, Wenyong Huang

## Abstract

**Purpose:** To compare macular thicknesses measured using swept-source optical coherence tomography (SS-OCT) and spectral domain OCT (SD-OCT) in normal subjects, diabetics with diabetic retinopathy (DR) and diabetics without DR (NDR).

**Methods:** We analysed 510 normal eyes, 741 NDR eyes and 209 DR eyes. Mean macular thicknesses in Early Treatment Diabetic Retinopathy Study (ETDRS) subfields, central point thicknesses (CPT), and macular volume were measured by SS-OCT and SD-OCT. We assessed agreement between SS-OCT and SD-OCT measurements by intraclass correlation coefficients (ICC) and Bland-Altman plots, and established a conversion equation relating central subfield (CSF), CPT and macular volume between the two devices.

**Results:** Macular thickness measurements by SS-OCT were significantly thinner than those by SD-OCT. The mean CSF thickness in normal eyes measured by SD-OCT and SS-OCT were 226.6 ± 19.1 μm (male 236.1 ± 19.1 μm vs female 223.0 ± 17.9 μm, p < 0.0001) and 258.4 ± 19.8 μm. In all three groups, the agreement between SS-OCT and SD-OCT was excellent (all ICC ≥ 0.866). For CSF the conversion equation SD-OCT = 31.95 + 0.999 × SS-OCT was derived. Using the equation, with 99.6% and 97.6% of the predicted values for CSF fell within 10% of the actual measurements in DR and NDR eyes, respectively.

**Conclusion:** We propose SS-OCT CSF thicknesses of 275 μm for males and 260 μm for females as the minimum criteria for macular edema in Chinese eyes. And SS-OCT measurements were significantly thinner than those of SD-OCT, we derived an equation to convert SS-OCT measurements to SD-OCT equivalents.

## INTRODUTION

Since its emergence in the 1990s,^1^ optical coherence tomography (OCT) has provided a non-invasive method for quantitative measurements retinal thickness and has become an important imaging technique for the evaluation and management of macular diseases. Macular thickness is one of the diagnostic criteria for macular diseases and central subfield (CSF) thickness measured using OCT are the most commonly used inclusion and retreatment criteria in clinical trials, such as diabetic macular edema (DME), age-related macular degeneration (AMD) and central retinal vein occlusion (CRVO).^2-5^ Spectral-domain OCT (SD-OCT), also known as Fourierdomain OCT, is currently the most widely used OCT technology, with fast scanning speed, short acquisition time and high-resolution to obtain retinal thickness.

Swept-source OCT (SS-OCT) is a new generation of OCT and has been introduced into clinical practice in recent years. This technology differs from conventional SD-OCT in that it, uses a light source with longer tunable wavelengths, has a more rapid scanning,^6, 7^ can penetrate deeper into ocular tissue, with little loss of signal, and allows for more detailed visualisation of retinal and choroidal structures.^8, 9^

In order to avoid misdiagnosis of macular disease in clinical work, ophthalmologist need to focus on the differences in macular thickness measured by various OCT devices. Some studies have compared macular thickness between the two methods and noted that the macular thickness measurements from SS-OCT were significantly thinner than those from SD-OCT,^25, 26^ which is due to the variation of the OCT segmentation algorithms that result in differences in the location of retinal inner boundary segmentation.^10-12^ SS-OCT as a relatively new imaging technique, normative database on macular thickness have not been established. It is thus important to compare these data with the commonly used SD-OCT data.

The purpose of the present study was to establish the normative data for macular thickness in Chinese eyes using the SS-OCT. Furthermore, we compared the macular thickness measurements from SS-OCT and SD-OCT, and sought to derive an equation to convert SS-OCT measurements to SD-OCT equivalents and evaluate the accuracy of the equation in the eyes of diabetics with diabetic retinopathy (DR) and those of diabetics without DR (NDR).

## METHODS

### Subjects

This prospective cross-sectional study was performed in the Zhongshan Ophthalmic Centre, which is affiliated with Sun Yat-sen University, Guangzhou, China. Subjects enrolled in the study between June 10, 2019 and December 22, 2019. All research conducted adhered to the tenets of the Declaration of Helsinki, and written informed consent was obtained from all subjects.

The inclusion criteria for normal subjects were as follows: best-corrected visual acuity of 20/20 or higher, intraocular pressure less than 21 mmHg, spherical refraction within ±6.0 dioptres (D), cylinder degree within ±3.0 D, normal fundus, and no history or evidence of diabetes, retinal disease, glaucoma, intraocular surgery or ocular laser treatment.

The inclusion criteria for diabetic patients were as follows: age of at least 30 years with type 2 diabetes, using standard seven-field Early Treatment Diabetic Retinopathy Study (ETDRS) photographs, the severity of diabetic retinopathy was assessed by an expert ophthalmologist (WW) according to the International Clinical Diabetic Retinopathy Disease Severity Scale,^13^ intraocular pressure less than 21 mmHg, spherical refraction within ±6.0D, cylinder degree within ±3.0D, no history of glaucoma, no retinal disease other than DR (such as epiretinal membrane, AMD, CRVO) and no history of previous intraocular treatment, including intravitreal injections, retinal laser procedures and intraocular surgery.

All subjects in this study underwent complete a ophthalmologic examination, including assessments of best-corrected visual acuity, intraocular pressure (IOP), refractive errors, slit-lamp biomicroscopy, fundus photography, axial length measurement (Lenstar LS900 Haag-Streit AG, Koeniz, Switzerland), SS-OCT and SD-OCT imaging, as well as a complete medical history.

### SS-OCT and SD-OCT imaging

All subjects underwent sequential scans with both SS-OCT and SD-OCT devices in the same clinical setting. The two OCT examinations were performed by the same experienced technician in the order of SD-OCT and SS-OCT, with each subject undergoing the second scan immediately after the first scan. SS-OCT (Triton DRI OCT, Topcon, Tokyo, Japan) uses a tunable laser as a light source to provide a 1050 nm centered wavelength, reaching a scanning speed of 100,000 A-scans per second and yielding 8 μm of axial and 20 μm of transverse resolution in tissues. A three-dimensional horizontal scan (7.0 mm × 7.0 mm, 512 × 256) protocol was adopted with centring on the fovea. SD-OCT (Spectralis OCT; Heidelberg Engineering, Heidelberg, Germany) use a super luminescence diode at the 870 nm wavelength as a light source; it provided 5-6 μm of axial resolution and 20 μm of transverse resolution with maximum scan speed of 40,000 A-scans per second. A 31 horizontal-line raster scan (30° × 25°, 20 mm × 20 mm) centred on the fovea was performed.

Images were excluded from analyses if the image quality score was below 15 dB for SD-OCT and below 45 for SS-OCT.^14,15^ Image quality scores were automatically generated by the software built into the OCT device. If images were decentration, they were also excluded during the image review process.

Retinal boundaries were automatically segmented by specialised image-viewing software from their respective OCT machines (Figure 1) and reviewed by trained retinal specialists (WW). In this study, segmentation errors of the eye were excluded. The mean macular thickness, central point thickness (CPT) and macular volume were automatically generated by the image-viewing software (Triton DRI OCT version 1.6.2.4, Heidelberg Eye Explorer version 6.0.9.0) in the ETDRS grid.

**Figure 1.**
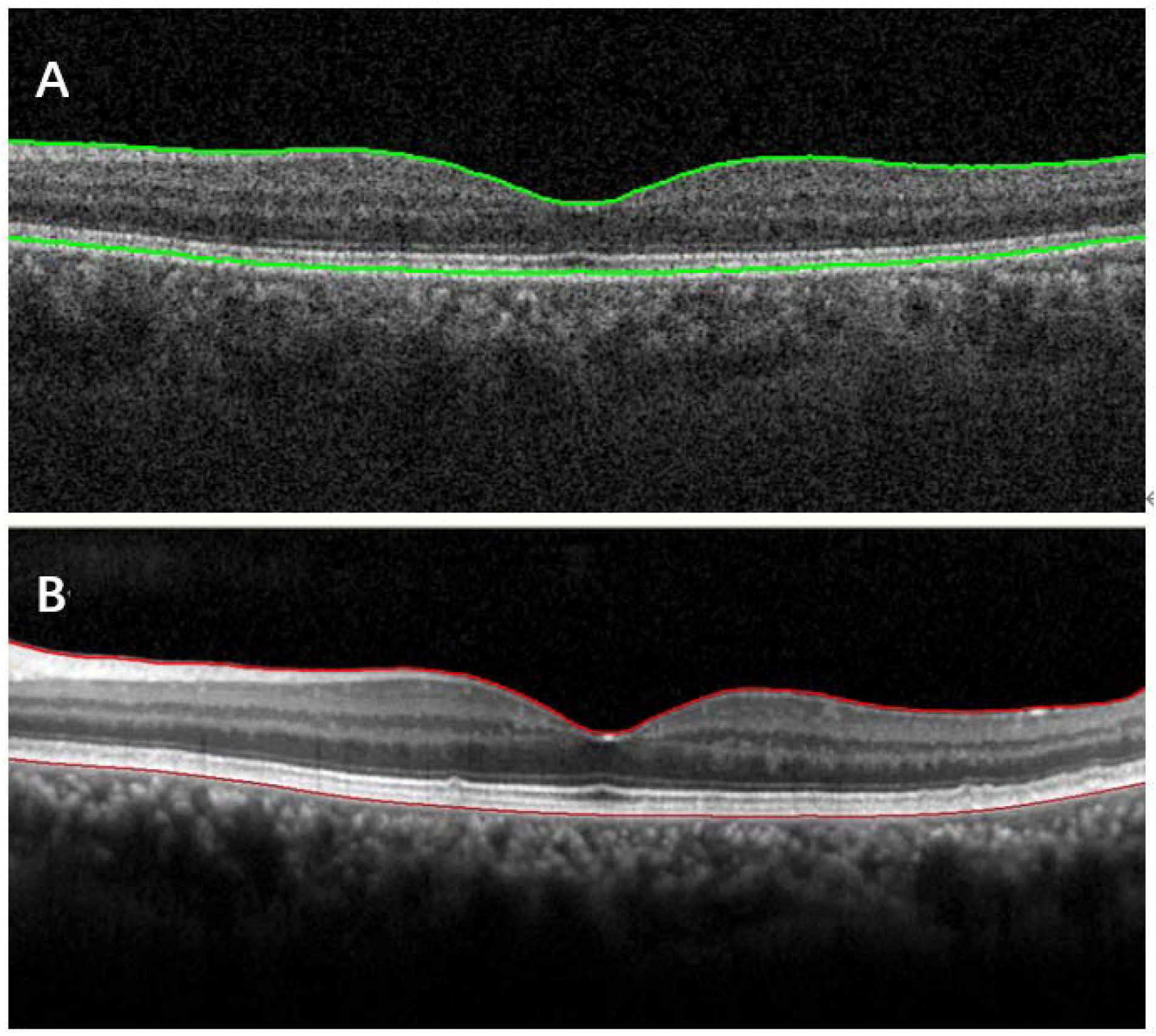
Retinal segmentation boundaries for SS-OCT and SD-CT. (A) Segmentation boundaries for SS-OCT, the inner segmentation boundary at the upper border of the retinal pigment epithelium (solid green line). (B) Segmentation boundaries for SD-OCT, the inner segmentation boundary at the retinal pigment epithelium-Bruch’s membrane complex (solid red line).

### Statistical analysis

Only data from the right eyes of subjects were included in the statistical analysis. Data are presented as mean values±standard deviations (SDs). Paired t-tests were used to compare macular thicknesses between SS-OCT and SD-OCT. Pearson correlation coefficients and intraclass correlation coefficients (ICC) were calculated to compare the correlation and agreement between the two OCT devices. The ICC was used to determine the agreement of two devices using a two-way random-effects model. The degree of reliability was classified into one the following five categories based on the ICC value: slight (0-0.2), fair (0.21-0.4), moderate (0.41-0.6), substantial (0.61-0.8), and almost perfect (0.81-1.0).^16^ The agreement between the two OCT devices was represented visually in Bland-Altman plots.

Conversion equation between SS-OCT and SD-OCT values was established according to the measurements of normal subjects. The conversion equation was derived from linear relationships of CSF, CPT and macular volume. The SD-OCT values in diabetic patients were predicted using the conversion equation based on the SS-OCT measurements and compared with the actual SD-OCT values. Bland-Altman plots were used to analyse the level of agreement between the predicted SD-OCT values and actual SD-OCT measurements. Statistical analyses were performed using SPSS version 25.0 (SPSS, Armonk, NY) and MedCalc version 16.8.4 (MedCalc Software bvba, Ostend, Belgium). A p-value less than 0.05 was considered to be statistically significant.

## RESULTS

### Demographics

A total of 1460 eyes from 1460 subjects were included in the study: 741 NDR eyes (mean age: 64.6 ± 7.6 years), 209 DR eyes (mean age: 64.3 ± 8.0 years), and 510 healthy eyes (mean age: 54.9 ± 9.1 years). There were significant differences in age, sex, spherical equivalent, and IOP among the three groups, but there were no significant differences in axial length. The study demographics and clinical characteristics are summarised in Table 1.

**Table 1.** Demographics and clinical characteristics of study patients.

|  | **Normal** | **NDR** | **DR** | **p Value** |
| --- | --- | --- | --- | --- |
| Gender (male/female) | 138 / 372 | 299 / 442 | 96 / 113 | <0.0001 |
| Age (years) | 54.9 ± 9.1 | 64.6 ± 7.6 | 64.3 ± 8.0 | <0.0001 |
| SE refraction (diopters) | 0.45 ± 1.96 | 0.96 ± 1.80 | 0.79 ± 1.84 | <0.0001 |
| Axial Length (mm) | 23.52 ± 0.99 | 23.49 ± 0.96 | 23.36 ± 0.98 | 0.118 |
| IOP (mm Hg) | 15.6 ± 2.1 | 16.1 ± 2.1 | 16.4 ± 2.4 | <0.0001 |

Data are expressed as number or mean ± standard deviation (SD).
SE = spherical equivalent; IOP = intraocular pressure.

### Thickness comparison between SS-OCT and SD-OCT

The mean macular thicknesses in nine ETDRS subfields, CPT and macular volume measured by SS-OCT and SD-OCT are shown in Table 2, Figure 2. There was significant differences in macular thickness measurements between SS-OCT and SD-OCT. The mean CSF thicknesses in normal eyes measured by SS-OCT and SD-OCT were 226.6 ± 19.1 μm and 258.4 ± 19.8 μm (p < 0.0001), respectively. There was a difference between genders, with mean SS-OCT CSF thickness being 236.1 ± 19.1 μm for males and 223.0 ± 17.9 μm for females (p < 0.0001); this difference was also seen for SD-OCT (267.6 ± 19.8 μm for males and 254.9 ± 18.6 μm for females, p < 0.0001). The mean differences in CSF thickness in NDR eyes and DR eyes were 32.1μm and 33.5 μm comparing SS-OCT and SD-OCT, respectively.

**Table 2.**
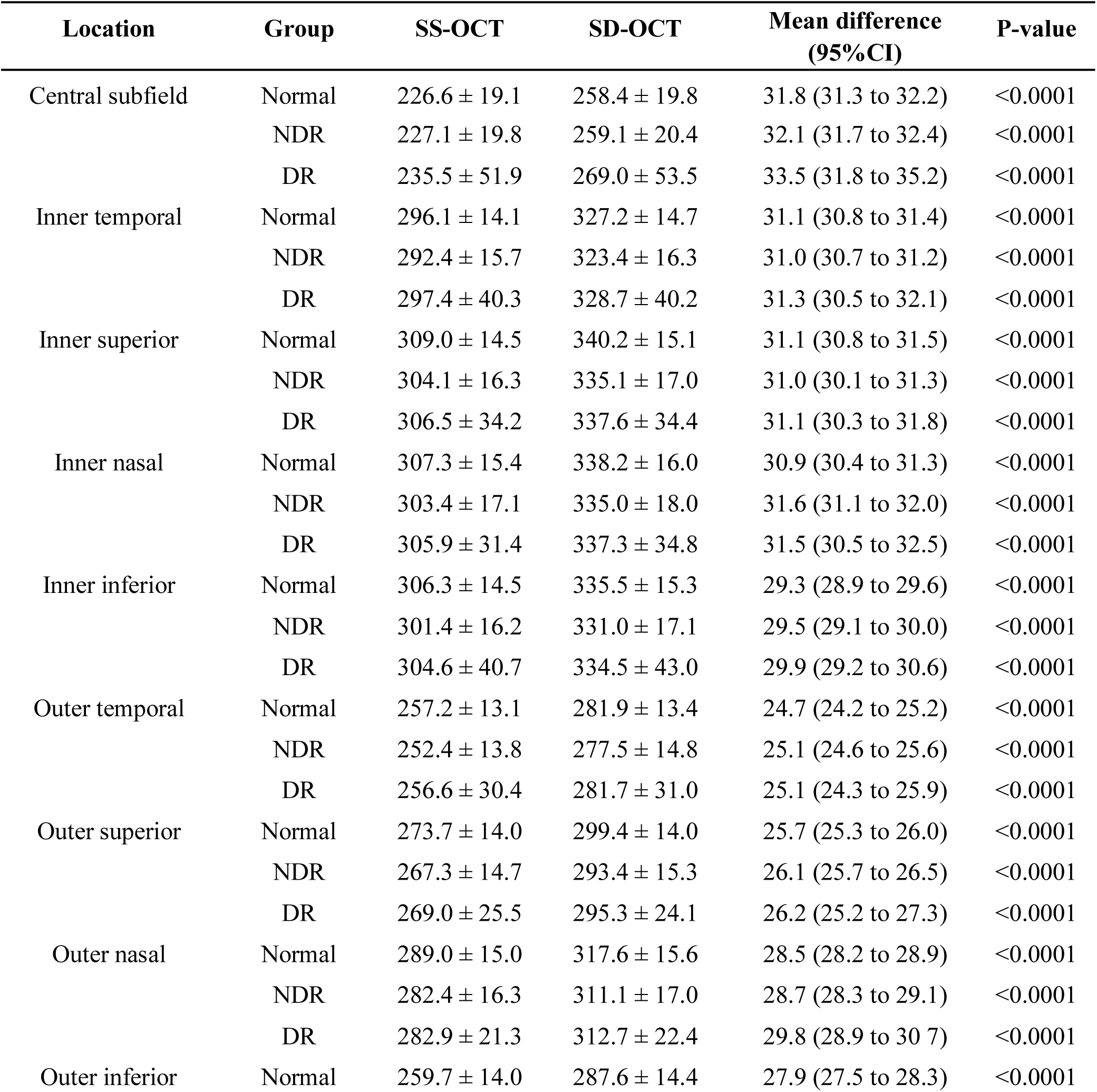
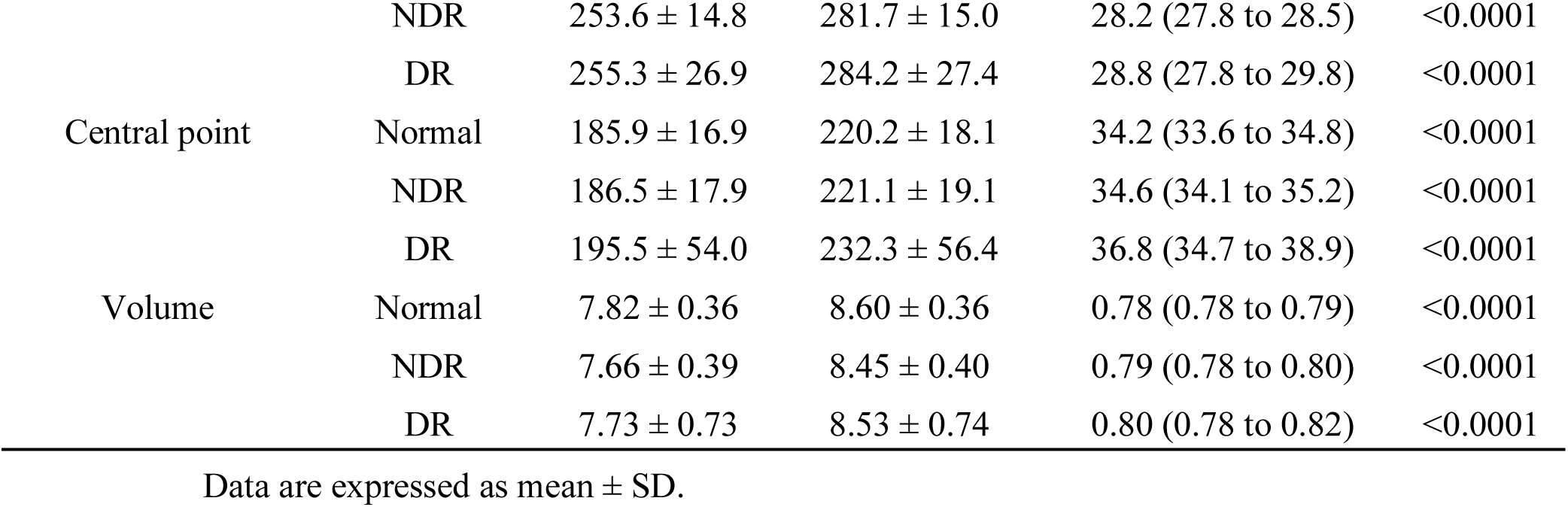
Comparison of macular thickness measurements between SS-OCT and SD-OCT

**Figure 2.**
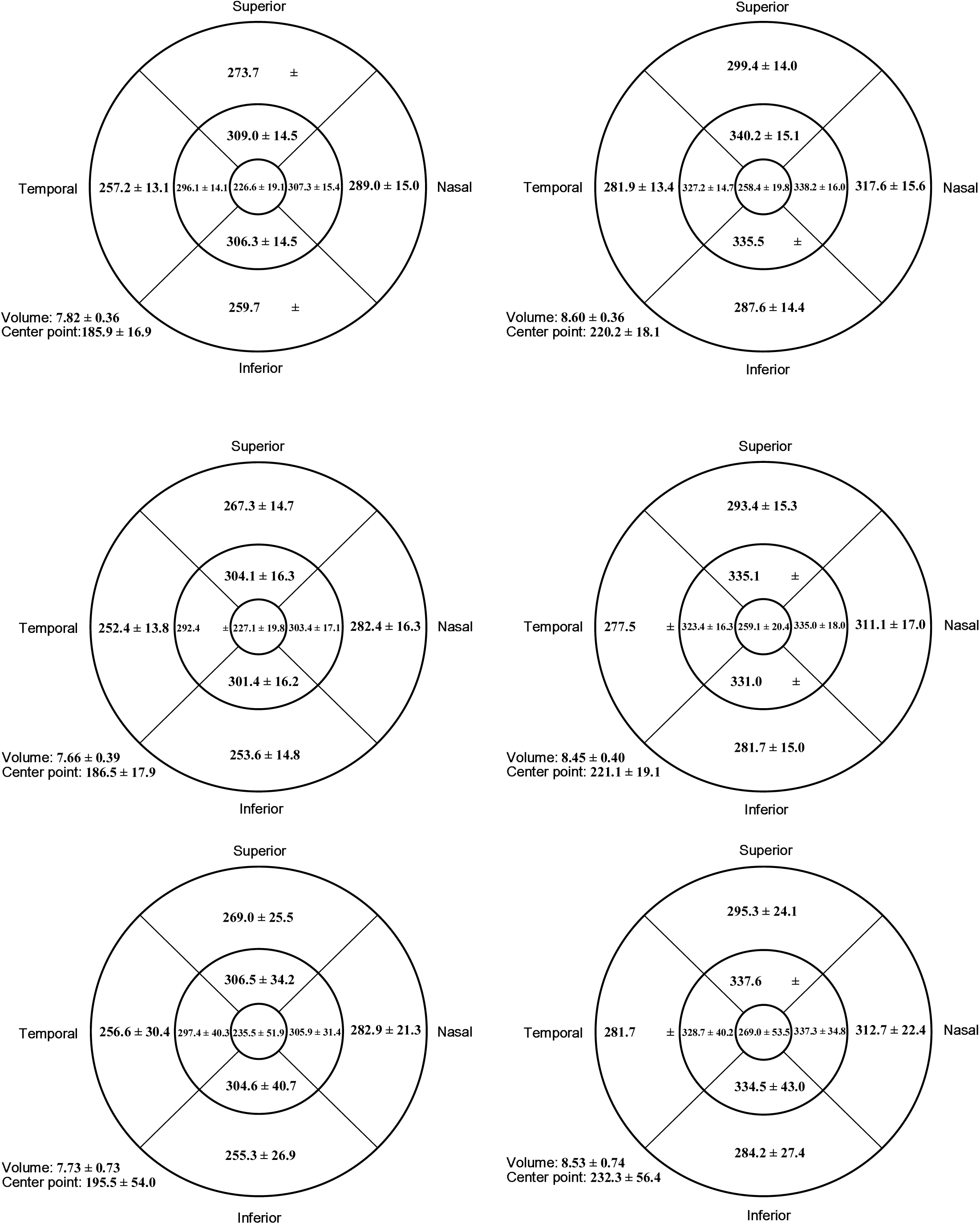
Mean macular thickness obtained in the nine ETDRS subfields on SS-OCT and SD-OCT. (A) SS-OCT macular thickness in normal subjects. (B) SD-OCT macular thickness in normal subjects. (C) SS-OCT macular thickness in NDR patients. (D) SD-OCT macular thickness in NDR patients. (E) SS-OCT macular thickness in DR patients. (F) SD-OCT macular thickness in DR patients.

### Correlation and agreement between SS-OCT and SD-OCT

SS-OCT and SD-OCT macular thicknesses were highly correlated (Table 3). The Pearson correlation coefficients for macular thickness in nine ETDRS subfields, CPT and macular volume were greater than 0.868 for all three groups of subjects. Likewise, all ICC values exceeded 0.866 (Table 3) and were classified as almost perfect (ICC = 0.81-1.0) for SS-OCT and SD-OCT, indicating the excellent degree of agreement of the measurements between the two OCT devices. The Bland-Altman plots shown in Figure 3 demonstrates good comparability of CSF thickness, CPT and macular volume measurements between SS-OCT and SD-OCT, with most of the values being within the limits of agreement.

**Table 3.**
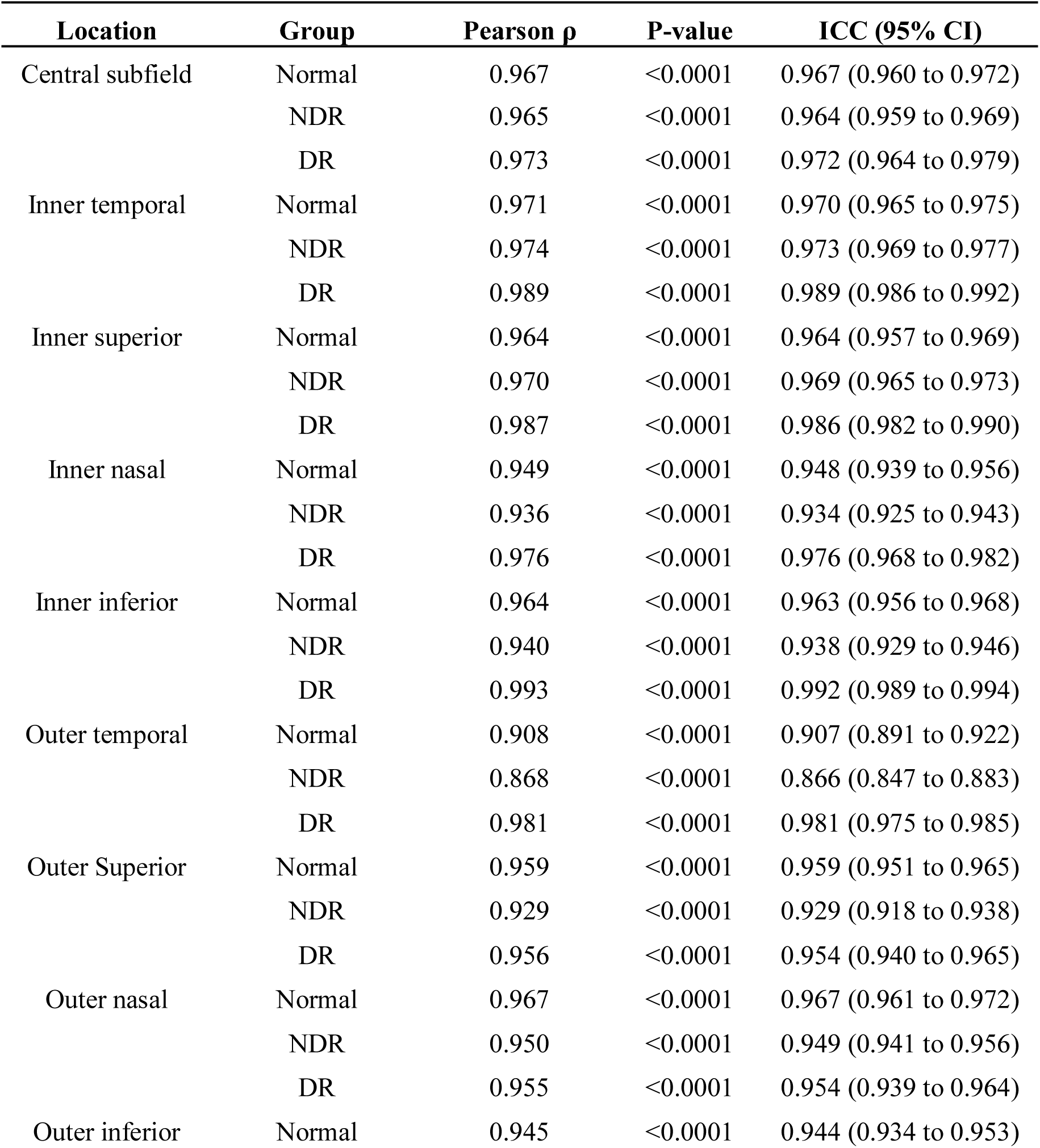
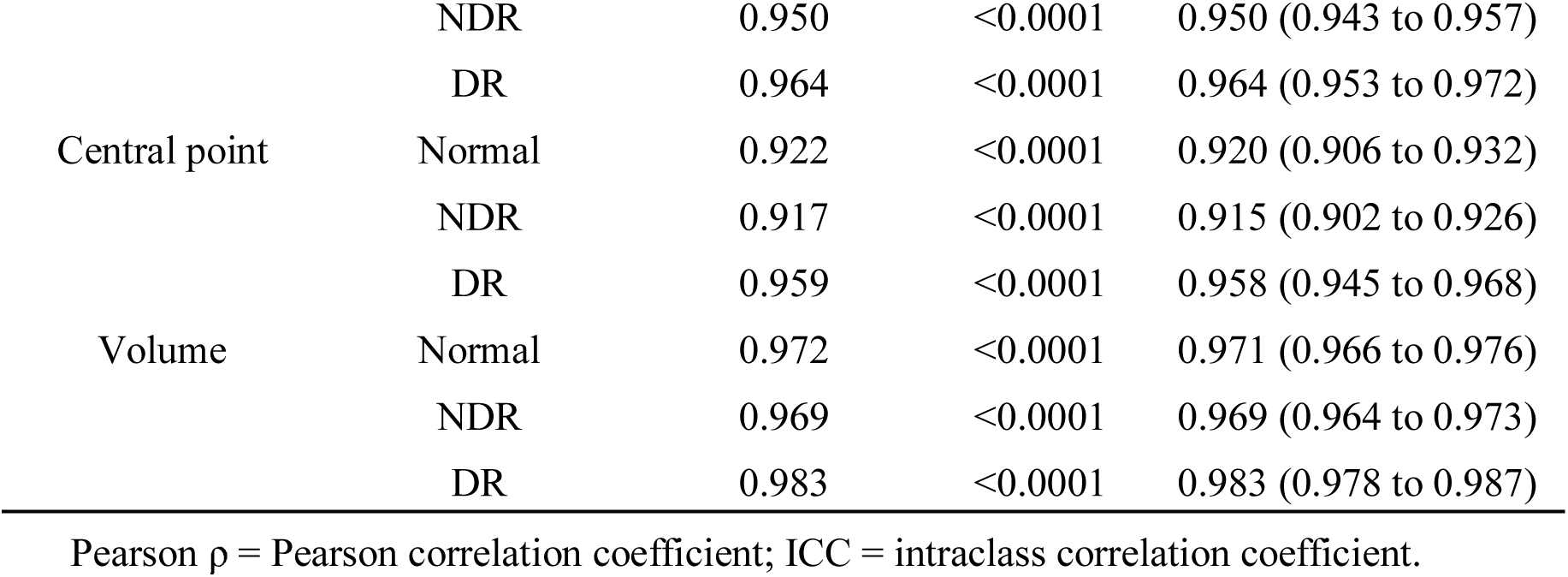
Correlation and agreement of macular thickness measurements for SS-OCT and SD-OCT

**Figure 3.**
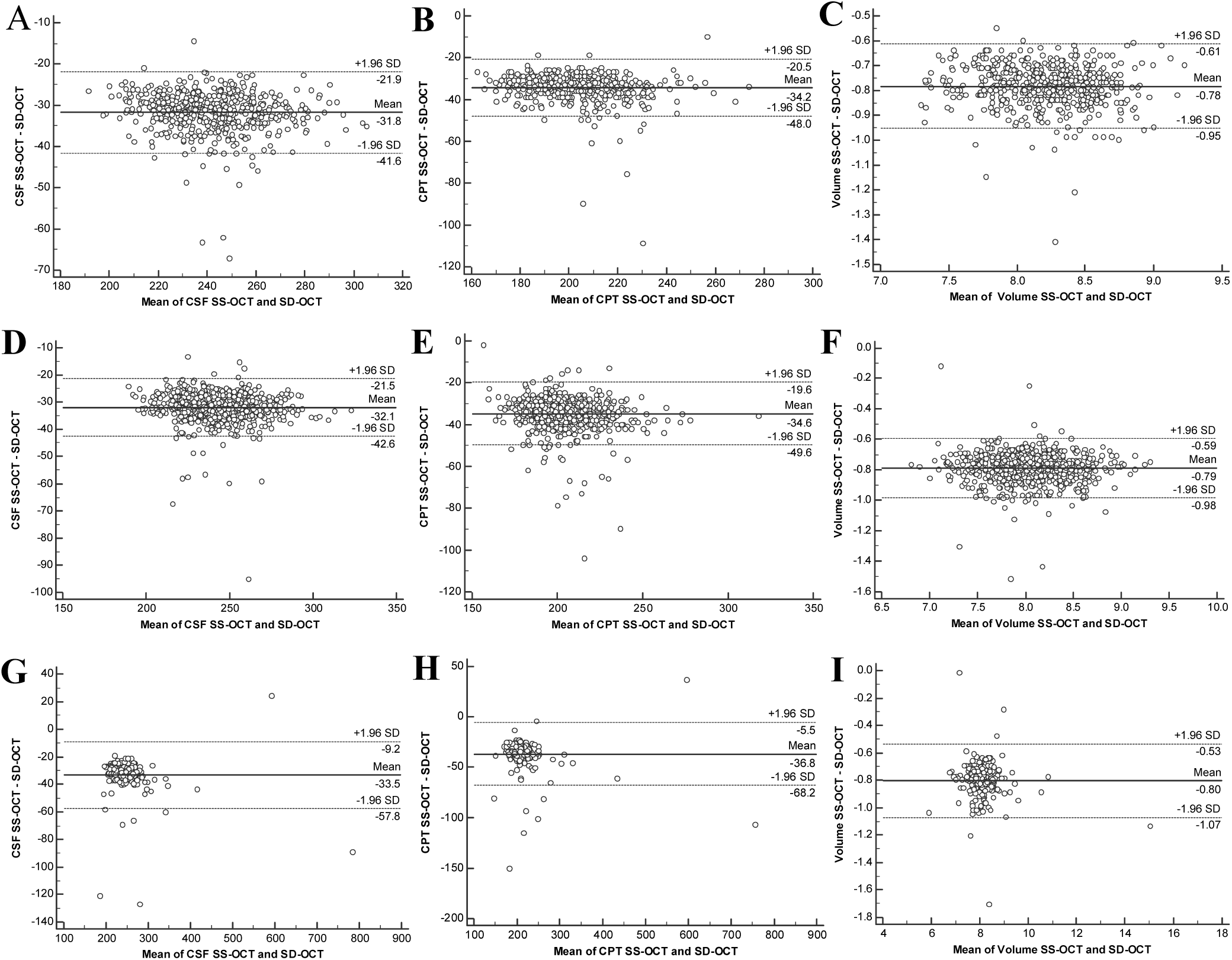
Bland-Altman plots of CSF, CPT and macular volume between SS-OCT and SD-OCT for normal subjects, NDR and DR patients. (A) CSF thickness in normal subjects. (B) CPT in normal subjects. (C) macular volume in normal subjects. (D) CSF thickness in NDR patients. (E) CPT in NDR patients. (F) macular volume in NDR patients. (G) CSF thickness in DR patients. (H) CPT in DR patients. (I) macular volume in DR patients.

### Conversion equation

The conversion equation was derived from measurements of normal eyes. The conversion equations for SS-OCT to SD-OCT are as follows: CSF SD-OCT = 31.95 + 0.999 × SS-OCT, CPT SD-OCT = 36.55 + 0.988 × SS-OCT, and macular volume SD-OCT = 0.84 + 0.993 × SS-OCT. The predicted SD-OCT values in NDR and DR eyes were compared with actual SD-OCT measurements (Table 4) with 99.6% and 97.6% of the predicted values for CSF falling within 10% of the actual measurements, and 98.5% and 95.2% of the predicted values for CSF falling within 5% of the actual measurements. The Bland-Altman plots in Figure 4 describes the level of agreement between predicted value and actual SD-OCT measurements.

**Table 4.**
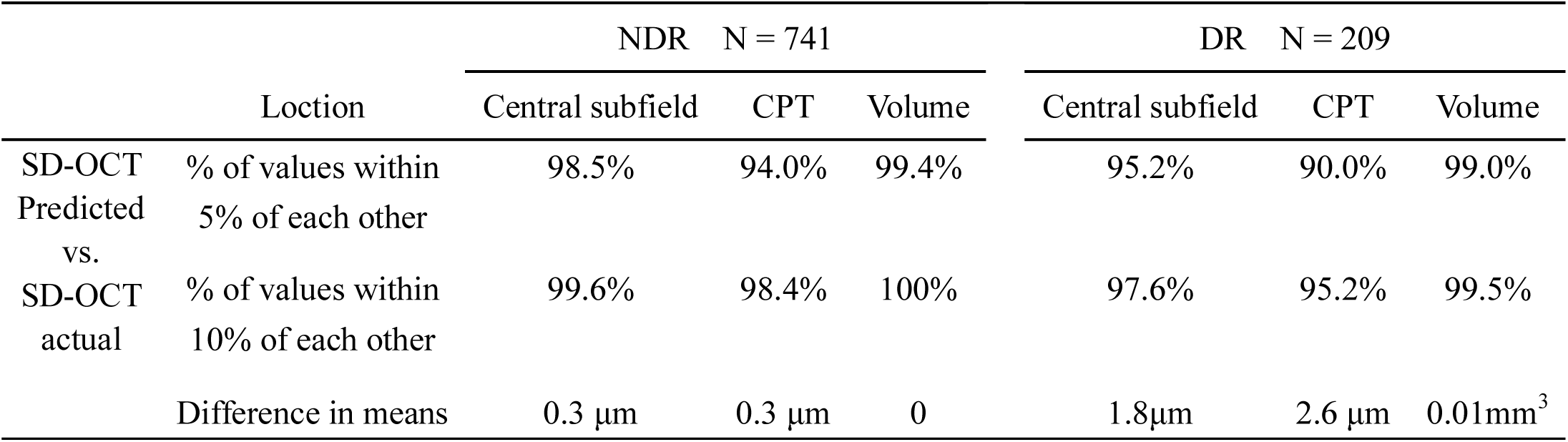
Conversion equation evaluation for SD-OCT: predicted values VS actual measurements.

**Figure 4.**
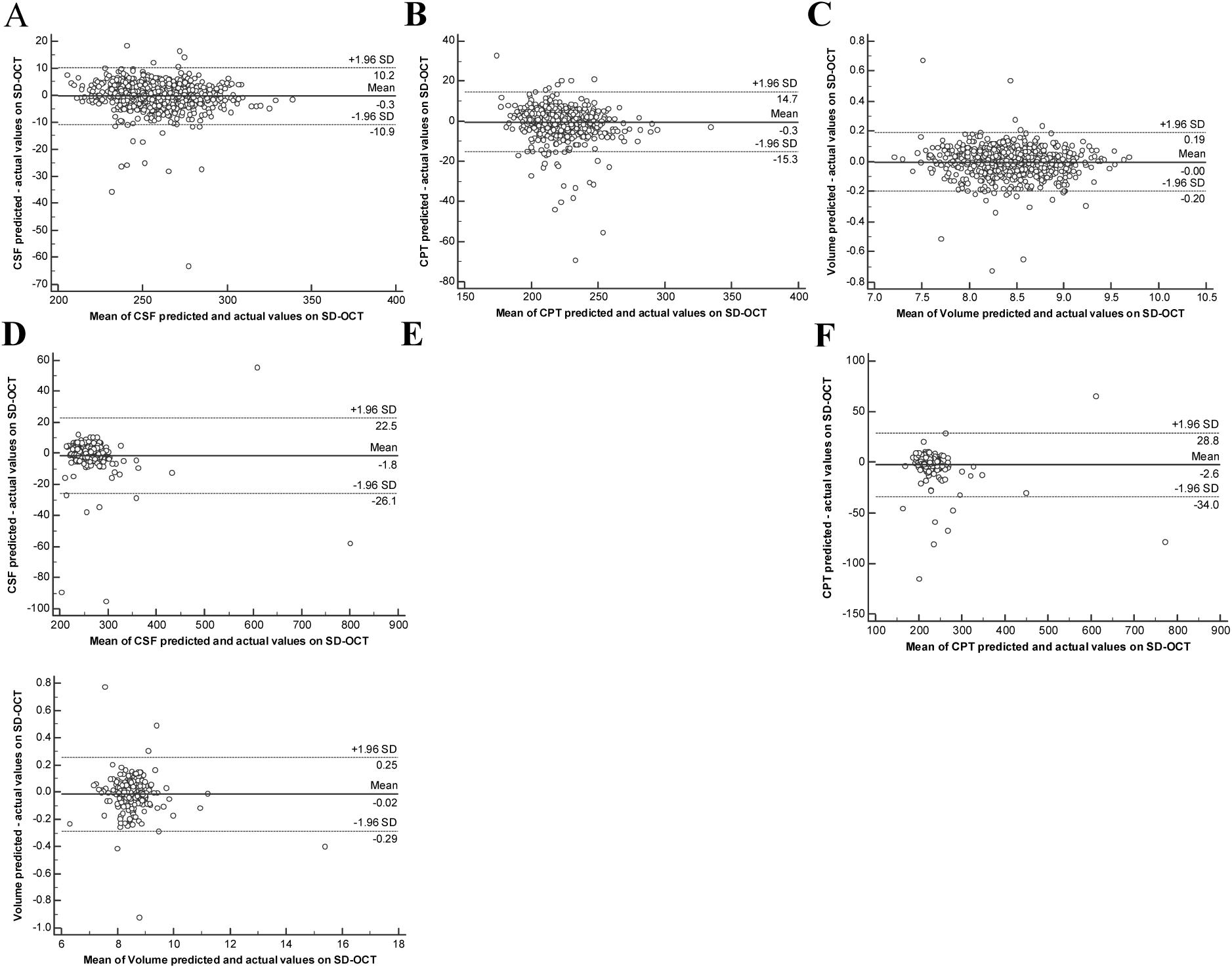
Bland-Altman plots showing the agreement between the predicted SD-OCT values in the CSF, CPT and macular volume and actual SD-OCT measurements. (A) CSF thickness in NDR patients. (B) CPT in NDR patients. (C) macular volume in DR patients. (D) CSF thickness in DR patients. (E) CPT in DR patients. (F) macular volume in NDR patients.

## DISCUSSION

We evaluated macualr thickness measurements from SS-OCT and SD-OCT devices in normal subjects, NDR patients and DR patients. The results revealed significant differences in macular thickness measurements between two OCT devices. The macular thicknesses measured using SS-OCT were significantly thinner in nine ETDRS subfields compared to those measured using commonly used SD-OCT. In addition, there was no difference in mean CSF thicknesses between normal eyes and NDR eyes, which was in parallel with previous studies.^17^

Central macular thickness is a common inclusion and retreatment criterion for macular pathology in clinical practice, such as DME, AMD and CRVO.^2-5^ Because the measured CSF thickness varies by the OCT technique used, the CSF thickness criterion for the diagnosis of macular disease also varies by OCT technique. For example, the CSF thicknesses cutoff point for Stratus and Spectral OCT diagnostic

DME are 250 μm and 315 μm, respectively.^18, 19^ As SD-OCT has been the most common commercially available OCT device over the past decade, most trials were designed based on the macular thickness measurements obtained from SD-OCT devices. We noted that our SS-OCT measurements were smaller than those of the SD-OCT, which may be due to differences in that segmentation of the inner retinal boundary resulting from the different segmentation software in each OCT device.^10-12^ Specifically, the inner boundary for SD-OCT is located at the retinal pigment epithelium (RPE)-Bruch’s membrane complex, whereas the inner boundary for SS-OCT is located at the upper border of the RPE (Figure 1).

Standardised reference values are essential for the diagnosis for macular edema. Our study showed that there were significant gender differences in CSF thickness as has been previously reported,^20, 21^ with the mean SS-OCT CSF thicknesses being 236.1 ± 19.1 μm and 223.0 ± 17.9 μm in healthy males and females, respectively. The diabetic retinopathy studies have defined a CSF of 250 μm as the upper limit of normal macular thickness using the Stratus OCT and this was calculated based on 2 SDs above the mean CSF.^18^ Similarly, in the study we propose CSF thicknesses of 275 μm for males and 260 μm for females as cut-off values for the presence of macular edema using SS-OCT in the Chinese population.

Although there are variations in the macular thickness values measured by different OCT devices, several earlier studies have investigated the agreement of macular thickness measurements between different OCT devices and shown that inter-device reproducibility was excellent.^22, 23^ However, fewer studies have compared SS-OCT and SD-OCT devices for the measurement of macular thickness. Tan et al.^24^ compared DRI-OCT-1 and Spectralis OCT in measuring the macular thicknesses of healthy eyes and eyes with high myopia; they found that the mean difference in CSF thickness was 33.1 μm and that the ICC value and Bland - Altman plots demonstrated excellent agreement between the two OCT devices. This result is similar to that of our study, in which the mean differences in CSF thickness measured using SS-OCT and SD-OCT in healthy subjects, NDR patients and DR patients were 31.8 μm, 32.1 μm and 33.5 μm, respectively. The Bland-Altman plots showed that most values were distributed close to the mean difference and were within the limits of agreement, which included 95% of the differences between measurements generated by the two devices. The Pearson correlation coefficient and ICC also indicated high correlation and excellent agreement in all sectors. However, Hanumunthad et al.^25^ reported that the mean DRI-OCT-1 CSF thickness in patients with AMD was 67.9 μm thinner than that obtained by Spectralis OCT, and that the Bland-Altman plot of the study showed low agreement in the macular thicknesses reported by the two OCT devices. The mean difference in CSF thickness was greater in patients with AMD, this may be attributed to the increased retinal thickness measured by SD-OCT, which may include drusen and other sub-RPE deposits.^26^

There are significant differences in macular thicknesses measured by different OCT methods, which introduces significant inconvenience to clinical work. Therefore, we have proposed to establish conversion equation between both types of OCT. Some studies have established equations to convert Stratus OCT CSF values to corresponding Cirrus, Spectralis, or RTVue OCT values,^27-29^ and noted relatively small differences between the predicted value and the actual measurements, indicating that the conversion equations were satisfactory. The Diabetic Retinopathy Clinical Research Network (DRCR.net) accepts up to a 20% change in CSF between two different OCT devices (after using the conversion equation) account for machine measurement errors and variations introduced by instrument alteration.^30^ In this study, we derived the conversion equation of SS-OCT to SD-OCT using measurements from normal eyes, using the conversion equation, with 99.6% and 97.6% of the predicted values for CSF fell within 10% of the actual measurements in DR and NDR eyes. Therefore, our equation can accurately predict the corresponding value of SD-OCT by using the SS-OCT measurements.

The strengths of this study include a larger sample size than those of previous studies, and the inclusion of only one eye per participant to rule out binocular interaction, which increased the reliability of the results. Additionally, this study made comparisons of macular thickness in all ETDRS subfields, CPT and macular volume using two commonly used OCT devices, not just the central subfield. Lastly, all OCT scans were obtained using standardised imaging protocols, and the two OCT scans were performed continuously to avoid the potential effects of diurnal variations.^31^

This study also has several limitations. First, in this study, SS-OCT and SD-OCT measurements were compared for the first time in Chinese eyes, and SS-OCT measurements of retinal thickness in normal eyes was reported, but it is known that macular thickness varies among ethnicities, with Asians and Blacks having thinner maculas compared to Whites.^32^ Therefore, more studies involving subjects of different ethnicities are needed to reach generalisable conclusions. Additionally, the difference in macular thickness measured by two OCT devices vary by the type of retinal disease.^26^ In other words, our equation can accurately predict SD-OCT values for the eyes of diabetic patients, but may not be applicable to other retinal diseases.

In conclusion, this study reports the differences in macular thicknesses measured by SS-OCT and SD-OCT in healthy subjects and diabetic patients with DR and without DR. We propose using SS-OCT CSF thicknesses of 275 μm for males and 260 μm for females as the minimum criteria for the presence of macular edema in Chinese eyes. Although the macular thickness measured by SS-OCT was significantly thinner than that of SD-OCT, there was high correlation and excellent agreement between the two OCT devices, which allowed a conversion equation to be established to compare the results of SS-OCT with those of SD-OCT.

## Data Availability

The availability of all data referred to in the manuscript and note links below.

## Author Contributions

WH and WW had full access to all the data in the study and take responsibility for the integrity of the data and the accuracy of the data analysis.

*Study concept and design:* WH, WW, KX, XG, WL, YL.

*Acquisition, analysis, or interpretation of data:* KX, WW, XG, WL, JM, YL.

*Drafting of the manuscript:* KX, WW.

*Critical revision of the manuscript for important intellectual content:*all authors.

*Statistical analysis:* KX.

*Administrative, technical, or material support:* WW, KX.

*Study supervision:* WW.

